## Supplementary material for "Asparagine metabolism in tumors is linked to poor survival in females with colorectal cancer: A cohort study": S1 Text, S1-S14 Table

**Supplementary materials**

**S1 Text. Timeline of sample collection and metabolite measurements and explanations**

In 2020, the clinical data on patient outcomes was retrieved from medical records held by Memorial Sloan-Kettering Cancer Center (MSKCC). All patients were followed from their operation date for follow up. The HILIC-MS analysis was performed from December 31, 2018, through January 4, 2019, and the RPLC-MS was performed from January 9, 2019, through January 18, 2019. Data analysis was conducted from March 25, 2020, through September 13, 2021.

The tissue samples were stored at -80 ℃ properly with minimum degradation that allowed us to perform metabolomics recently, and thus to find associations between metabolites and prognostic outcomes. The patients were not treated with chemotherapy before the samples were taken. In addition, the chemotherapy provided was standardized between patients, which hasn’t changed considerably since then for stage I-III patients, so as to provide us a pretherapeutic metabolic profile.

**S1 Table. Demographic characteristics and clinical factors for each subgroup.**

| **Subgroup** | **Stage I (n=47)** | | **Stage II (n=86)** | | **Stage III (n=64)** | |
| --- | --- | --- | --- | --- | --- | --- |
|  | **RCC**  **(n=22)** | **LCC**  **(n=25)** | **RCC**  **(n=44)** | **LCC**  **(n=42)** | **RCC**  **(n=32)** | **LCC**  **(n=32)** |
| **Sex, n** | | | | | | |
| Males | 10 | 15 | 23 | 25 | 15 | 14 |
| Females | 12 | 10 | 21 | 17 | 17 | 18 |
| **Age, y, mean (SD)** | | | | | | |
| Males | 73.9 (6.5) | 69.3 (5.8) | 72.9 (7.8) | 72.2 (8.5) | 73.5 (7.8) | 63.7 (5.8) |
| Females | 72.1 (6.2) | 69.6 (7.6) | 73.5 (9.8) | 69.1 (7.8) | 72.2 (6.6) | 71.1 (6.0) |
| **Race/Ethnicity, n** | | | | | | |
| NHWs | 20 | 22 | 40 | 37 | 26 | 25 |
| Hispanic | 2 | 3 | 2 | 3 | 4 | 2 |
| AA | 0 | 0 | 1 | 0 | 1 | 4 |
| API | 0 | 0 | 1 | 2 | 1 | 1 |
| **5**-**year Overall survival rate, %^a^** | | | | | | |
| Males | 87.5 | 85.1 | 76.5 | 74.1 | 47.1 | 78.6 |
| Females | 100 | 100 | 82.9 | 100 | 65.2 | 61.8 |
| **5**-**year Recurrence**-**free survival rate, %^a^** | | | | | | |
| Males | 90.0 | 78.3 | 86.8 | 67.6 | 82.1 | 70.1 |
| Females | 90.9 | 100.0 | 87.5 | 87.4 | 76.0 | 68.8 |

RCC: right-sided colon cancer, LCC: left-sided colon cancer, NHWs: non-Hispanic whites, AA: African-Americans, API: Asian-Pacific Islander

^a^ The survival rates were calculated using the Kaplan-Meier estimation method.

**S2 Table.** Prognosis of patients in early stage (Stage I and II) vs late stage (Stage III)

| **5**-**year Overall Survival (OS)** | | | | | | | | | | | | | | | |
| --- | --- | --- | --- | --- | --- | --- | --- | --- | --- | --- | --- | --- | --- | --- | --- |
| **Subgroup** | **RCC males** | | | | **RCC females** | | | | **LCC males** | | | | **LCC females** | | |
| **Event^a^** | 0 | | 1 | | 0 | | 1 | | 0 | | 1 | | 0 | | 1 |
| **Early stage^b^** | 28 | | 5 | | 30 | | 3 | | 32 | | 8 | | 27 | | 0 |
| **Late stage^c^** | 8 | | 7 | | 12 | | 5 | | 11 | | 3 | | 12 | | 6 |
| **Total** | 36 | | 12 | | 42 | | 8 | | 43 | | 11 | | 39 | | 6 |
| **5**-**year Recurrence**-**free Survival (RFS)** | | | | | | | | | | | | | | | |
| **Subgroup** | **RCC males** | | | **RCC females** | | | | **LCC males** | | | | **LCC females** | | | |
| **Event^d^** | 0 | 1 | | 0 | | 1 | | 0 | | 1 | | 0 | | 1 | |
| **Early stage^b^** | 30 | 3 | | 30 | | 3 | | 32 | | 8 | | 25 | | 2 | |
| **Late stage^c^** | 13 | 2 | | 14 | | 3 | | 10 | | 4 | | 13 | | 5 | |
| **Total** | 43 | 5 | | 44 | | 6 | | 42 | | 12 | | 38 | | 7 | |

^a^ Event = death. Event =1 represents that death occurred within 5 years of follow-up, and event = 0 indicates the event did not occur within 5 years of follow-up (survived or scored as censored).

^b^ Early stage combines stage I and II together.

^c^ Late stage refers to stage III.

^d^ Event = recurrence. Event = 1 indicates that recurrence occurred within 5 years of follow-up, event = 0 indicates that recurrence did not occur within 5 years of follow-up. If a patient died before CRC recurrence occurred, it is counted towards a death event.

Notes:

If a patient died for any reason without recurrence within 5 years of follow-up, it is only counted toward a death event in OS.

If a patient experienced recurrence but did not die within 5 years of follow-up, it is only counted toward a recurrence event in RFS.

If a patient experienced recurrence and then died within 5 years of follow-up, it is counted toward a recurrence event in RFS and a death event in OS.

If a patient experienced neither recurrence nor death within 5 years of follow-up, the event is 0 for both OS and RFS.

Due to the absence of death events in females of clinical stage I, we regarded stage I and II as early stage, and stage III as late stage.

**S3 Table**. Multivariate analyses of the associations between individual metabolites and 5-year OS by sex, adjusted for anatomic location, chemotherapy history, clinical stages, and age.

| **Metabolite name** | **Females** | | | **Males** | | | **Int. Sex *P* value^b^** |
| --- | --- | --- | --- | --- | --- | --- | --- |
|  | **HR** | **95% CI** | ***P* value^a^** | **HR** | **95% CI** | ***P* value^a^** |  |
| Acetyl-lysine | 0.97 | 0.75-1.25 | 0.809 | 0.82 | 0.71-0.96 | 0.012 | 0.298 |
| Adenosine | 0.90 | 0.69-1.17 | 0.421 | 1.29 | 1.03-1.62 | 0.026 | 0.038 |
| Alanine | 1.06 | 0.78-1.44 | 0.705 | 0.77 | 0.61-0.98 | 0.034 | 0.092 |
| Argininosuccinic acid | 0.93 | 0.71-1.23 | 0.629 | 0.74 | 0.58-0.93 | 0.010 | 0.151 |
| Asparagine | 1.51 | 0.89-2.57 | 0.127 | 0.72 | 0.54-0.96 | 0.025 | 0.026 |
| Carnitine | 0.61 | 0.04-9.10 | 0.717 | 0.53 | 0.30-0.94 | 0.031 | 0.897 |
| Citrulline | 1.66 | 0.97-2.83 | 0.065 | 0.65 | 0.46-0.92 | 0.014 | 0.002 |
| Glycerol 3-phosphate | 3.64 | 1.26-10.49 | 0.017 | 0.91 | 0.47-1.77 | 0.777 | 0.018 |
| Hypoxanthine | 1.05 | 0.35-3.14 | 0.929 | 0.65 | 0.44-0.95 | 0.027 | 0.441 |
| LysoPC(16:0) | 1.52 | 1.03-2.25 | 0.035 | 0.85 | 0.65-1.11 | 0.242 | 0.009 |
| Ornithine | 0.97 | 0.56-1.66 | 0.904 | 0.68 | 0.47-0.97 | 0.035 | 0.310 |
| Serine | 1.25 | 0.65-2.41 | 0.497 | 0.55 | 0.37-0.81 | 0.002 | 0.035 |
| Spermine | 1.49 | 1.04-2.13 | 0.031 | 1.03 | 0.83-1.27 | 0.811 | 0.073 |
| Succinate | 0.35 | 0.12-0.99 | 0.047 | 1.91 | 1.23-2.96 | 0.004 | 0.004 |
| Threonine | 1.12 | 0.67-1.87 | 0.670 | 0.61 | 0.44-0.85 | 0.004 | 0.035 |
| UDP-D-Glucose | 0.81 | 0.67-0.97 | 0.025 | 1.16 | 0.95-1.41 | 0.152 | 0.010 |
| Uracil | 1.24 | 0.56-2.76 | 0.599 | 0.42 | 0.26-0.69 | 0.001 | 0.024 |
| Xanthosine | 1.22 | 0.86-1.73 | 0.266 | 0.71 | 0.54-0.94 | 0.016 | 0.028 |

^a^ *P* values for the association between individual metabolites and 5-year OS calculated by multivariate Cox PH regression models, adjusted for anatomic location, clinical stages, chemotherapy history and age (before FDR adjustment). The abundance of each metabolite was treated as a continuous variable and was log_2_ transformed. A metabolite with HR < 1 was associated with better OS; a metabolite with HR > 1 was associated with worse OS.

^b^ *P* value of the interaction between the metabolite and sex. Int. Sex *P* value < .05 indicate a significant sex interaction.

**S4 Table**. Multivariate analyses of the associations between individual metabolites and 5-year RFS by sex, adjusted for anatomic location, chemotherapy history, clinical stages, and age.

| **Metabolite name** | **Females** | | | **Males** | | | **Int. Sex *P* value^b^** |
| --- | --- | --- | --- | --- | --- | --- | --- |
|  | **HR** | **95% CI** | ***P* value^a^** | **HR** | **95% CI** | ***P* value^a^** |  |
| Acetyl-lysine | 0.98 | 0.76-1.26 | 0.855 | 0.78 | 0.67-0.92 | 0.002 | 0.076 |
| Alanine | 1.12 | 0.81-1.55 | 0.495 | 0.74 | 0.58-0.94 | 0.013 | 0.053 |
| AMP | 1.10 | 0.69-1.77 | 0.689 | 0.69 | 0.51-0.95 | 0.021 | 0.124 |
| Argininosuccinic acid | 1.03 | 0.79-1.35 | 0.825 | 0.70 | 0.54-0.92 | 0.011 | 0.036 |
| Asparagine | 1.54 | 0.93-2.56 | 0.093 | 0.74 | 0.56-0.97 | 0.031 | 0.007 |
| Carnitine | 3.52 | 0.25-50.41 | 0.354 | 0.37 | 0.20-0.66 | 0.001 | 0.117 |
| CMP | 0.97 | 0.57-1.65 | 0.906 | 0.70 | 0.50-0.96 | 0.029 | 0.287 |
| Creatinine | 0.75 | 0.42-1.32 | 0.317 | 1.70 | 1.15-2.51 | 0.008 | 0.040 |
| Cytidine | 0.46 | 0.28-0.75 | 0.002 | 0.75 | 0.49-1.16 | 0.194 | 0.134 |
| Fructose 6-phosphate | 0.84 | 0.55-1.28 | 0.414 | 0.68 | 0.49-0.95 | 0.026 | 0.534 |
| Glutamine | 0.77 | 0.26-2.30 | 0.642 | 0.47 | 0.23-0.97 | 0.041 | 0.338 |
| Glutathione | 0.73 | 0.56-0.95 | 0.019 | 0.92 | 0.79-1.06 | 0.246 | 0.302 |
| Glutathione disulfide | 0.72 | 0.57-0.91 | 0.006 | 0.81 | 0.67-0.98 | 0.034 | 0.092 |
| GMP | 1.04 | 0.69-1.58 | 0.838 | 0.74 | 0.57-0.96 | 0.024 | 0.169 |
| Hypoxanthine | 1.95 | 0.59-6.45 | 0.276 | 0.32 | 0.20-0.51 | <0.001 | 0.008 |
| LysoPC(16:1) | 1.06 | 0.76-1.49 | 0.729 | 0.77 | 0.61-0.97 | 0.025 | 0.106 |
| LysoPE(18:2) | 1.07 | 0.70-1.65 | 0.750 | 0.72 | 0.55-0.96 | 0.023 | 0.127 |
| LysoPE(20:1) | 0.98 | 0.77-1.24 | 0.856 | 0.83 | 0.70-0.98 | 0.031 | 0.225 |
| LysoPE(22:5) | 1.09 | 0.70-1.70 | 0.701 | 0.70 | 0.53-0.92 | 0.010 | 0.072 |
| Serine | 1.45 | 0.74-2.84 | 0.282 | 0.57 | 0.40-0.83 | 0.003 | 0.010 |
| Sphinganine-1-phosphate | 0.94 | 0.60-1.47 | 0.777 | 0.66 | 0.45-0.95 | 0.026 | 0.216 |
| Stearamide | 0.92 | 0.63-1.36 | 0.674 | 0.70 | 0.53-0.92 | 0.012 | 0.211 |
| Threonine | 0.98 | 0.58-1.64 | 0.929 | 0.64 | 0.46-0.89 | 0.007 | 0.074 |
| Xanthine | 0.62 | 0.38-1.01 | 0.053 | 0.69 | 0.48-0.98 | 0.036 | 0.869 |
| Xanthosine | 1.01 | 0.72-1.40 | 0.973 | 0.71 | 0.52-0.98 | 0.040 | 0.112 |

^a^ *P* values for the association between individual metabolites and 5-year RFS calculated by multivariate Cox PH regression models, adjusted for anatomic location, clinical stages, chemotherapy history and age (before FDR adjustment). The abundance of each metabolite was treated as a continuous variable and was log_2_ transformed. A metabolite with HR < 1 was associated with better RFS; a metabolite with HR > 1 was associated with worse RFS.

^b^ *P* value of the interaction between the metabolite and sex. Int. Sex *P* value < .05 indicate a significant sex interaction.

**S5 Table.** Multivariate analysis of association between metabolites in ASNS-catalyzed asparagine synthesis pathway and OS and RFS for all patients

| **Variable^a^** | **OS** | | | | **RFS** | | | |
| --- | --- | --- | --- | --- | --- | --- | --- | --- |
|  | **HR** | **95% CI** | ***P* value** | **Int. Sex *P* value^b^** | **HR** | **95% CI** | ***P* value** | **Int. Sex *P* value^b^** |
| Asparagine | 0.89 | 0.61-1.29 | 0.535 | 0.021 | 1.27 | 0.84-1.92 | 0.251 | 0.003 |
| Aspartate | 1.25 | 0.85-1.84 | 0.257 | 0.639 | 1.31 | 0.86-2.00 | 0.202 | 0.237 |
| Glutamate | 0.93 | 0.45-1.90 | 0.837 | 0.808 | 0.93 | 0.44-1.97 | 0.841 | 0.064 |
| Glutamine | 0.86 | 0.35-2.08 | 0.734 | 0.974 | 0.37 | 0.16-0.87 | 0.023 | 0.285 |
| AMP | 0.83 | 0.64-1.06 | 0.135 | 0.860 | 0.75 | 0.57-0.98 | 0.034 | 0.295 |
| Sex = Male (ref: Female) | 2.01 | 1.00-4.05 | 0.051 | - | 1.12 | 0.53-2.38 | 0.761 | - |
| Anatomic location = RCC (ref: LCC) | 0.87 | 0.43-1.77 | 0.710 | - | 0.67 | 0.31-1.44 | 0.300 | - |
| Clinical stage = late (ref: early) | 4.48 | 2.26-9.24 | 0.003 | - | 1.91 | 0.63-5.79 | 0.252 | - |
| Chemotherapy = Yes (ref: No) | 1.03 | 0.38-2.76 | 0.956 |  | 1.26 | 0.41-3.86 | 0.685 |  |
| Age | 1.10 | 1.05-1.15 | <0.001 | - | 0.97 | 0.92-1.03 | 0.304 | - |

^a^ All the variables listed were included in one multivariate Cox PH model where the abundance of each metabolite was treated as a continuous variable and was log_2_ transformed.

^b^ *P* value of the interaction between the metabolite and sex.

**S6 Table**. Multivariate analysis of association between metabolites in ASNS-catalyzed asparagine synthesis metabolic pathway and OS and RFS by sex

| **Variable^a^** | **OS** | | | | | | **RFS** | | | | | |
| --- | --- | --- | --- | --- | --- | --- | --- | --- | --- | --- | --- | --- |
|  | **Females** | | | **Males** | | | **Females** | | | **Males** | | |
|  | **HR** | **95% CI** | ***P* value** | **HR** | **95% CI** | ***P* value** | **HR** | **95% CI** | ***P* value** | **HR** | **95% CI** | ***P* value** |
| Asparagine | 6.39 | 1.78-22.91 | 0.004 | 0.57 | 0.36-0.91 | 0.018 | 4.36 | 1.39-13.68 | 0.012 | 0.96 | 0.61-1.50 | 0.856 |
| Aspartate | 1.47 | 0.64-3.40 | 0.365 | 1.32 | 0.75-2.32 | 0.339 | 1.60 | 0.71-4.05 | 0.239 | 1.40 | 0.79-2.50 | 0.252 |
| Glutamate | 0.24 | 0.06-0.95 | 0.042 | 1.28 | 0.48-3.38 | 0.618 | 0.76 | 0.19-3.11 | 0.705 | 0.75 | 0.28-2.02 | 0.573 |
| Glutamine | 0.16 | 0.03-1.06 | 0.057 | 1.18 | 0.39-3.54 | 0.772 | 0.10 | 0.02-0.57 | 0.010 | 0.52 | 0.17-1.63 | 0.266 |
| AMP | 0.65 | 0.31-1.37 | 0.252 | 0.89 | 0.65-1.22 | 0.476 | 0.59 | 0.25-1.36 | 0.214 | 0.70 | 0.49-1.00 | 0.049 |
| Anatomic location = RCC (ref: LCC) | 1.63 | 0.47-5.65 | 0.438 | 0.80 | 0.31-2.10 | 0.653 | 0.78 | 0.23-2.65 | 0.692 | 0.61 | 0.20-1.84 | 0.380 |
| Clinical stage = late (ref: early) | 14.90 | 2.19-101.43 | 0.006 | 6.02 | 1.39-26.06 | 0.016 | 1.92 | 0.34-10.99 | 0.462 | 1.29 | 0.23-7.09 | 0.771 |
| Chemotherapy = Yes (ref: No) | 1.78 | 0.39-8.23 | 0.458 | 0.74 | 0.18-2.99 | 0.676 | 2.34 | 0.38-14.29 | 0.358 | 1.01 | 0.19-5.24 | 0.991 |
| Age | 1.09 | 1.00-1.19 | 0.037 | 1.15 | 1.07-1.23 | <0.001 | 1.01 | 0.91-1.11 | 0.905 | 0.94 | 0.87-1.01 | 0.112 |

^a^ All the variables listed were included in one multivariate Cox PH model. The abundance of each metabolite was treated as a continuous variable and was log_2_ transformed.

**S7 Table.** Multivariate analysis of association between metabolites in PPP and glycolysis metabolic pathway and OS and RFS for all patients

| **Variable^a^** | **OS** | | | | **RFS** | | | |
| --- | --- | --- | --- | --- | --- | --- | --- | --- |
|  | **HR** | **95% CI** | ***P* value^a^** | **Int. Sex *P* value^b^** | **HR** | **95% CI** | ***P* value^a^** | **Int. Sex *P* value^b^** |
| Ribulose 5-phosphate | 1.18 | 0.84-1.65 | 0.354 | 0.934 | 1.33 | 0.89-1.99 | 0.164 | 0.263 |
| Lactate | 1.49 | 0.81-2.75 | 0.203 | 0.601 | 1.21 | 0.51-2.85 | 0.660 | 0.472 |
| DHAPorG3P | 1.21 | 0.72-2.03 | 0.479 | 0.214 | 0.99 | 0.57-1.71 | 0.975 | 0.137 |
| Glucose 6-phosphate | 0.95 | 0.67-1.35 | 0.775 | 0.817 | 1.30 | 0.90-1.87 | 0.156 | 0.110 |
| Fructose 6-phosphate | 0.68 | 0.42-1.10 | 0.114 | 0.899 | 0.46 | 0.27-0.78 | 0.004 | 0.615 |
| Glycerol 3-phosphate | 1.44 | 0.80-2.59 | 0.222 | 0.046 | 0.53 | 0.25-1.12 | 0.097 | 0.253 |
| Phosphoenolpyruvate | 1.05 | 0.83-1.35 | 0.670 | 0.241 | 1.17 | 0.84-1.61 | 0.358 | 0.418 |
| Sex = Male (ref: Female) | 1.95 | 0.97-3.90 | 0.060 | - | 1.33 | 0.64-2.78 | 0.442 | - |
| Anatomic location = RCC (ref: LCC) | 0.65 | 0.29-1.42 | 0.279 | - | 0.83 | 0.36-1.93 | 0.665 | - |
| Clinical stage = late (ref: early) | 4.24 | 1.58-11.42 | 0.004 | - | 1.53 | 0.46-5.14 | 0.491 | - |
| Chemotherapy = Yes (ref: No) | 1.01 | 0.36-2.85 | 0.980 | - | 1.63 | 0.47-5.72 | 0.442 | - |
| Age | 1.11 | 1.06-1.16 | <0.001 | - | 0.99 | 0.93-1.04 | 0.596 | - |

^a^ All the variables listed were included in one multivariate Cox PH model where the abundance of each metabolite was treated as a continuous variable and was log_2_ transformed.

^b^ *P* value of the interaction between the metabolite and sex.

**S8 Table.** Multivariate analysis of association between metabolites in PPP and glycolysis metabolic pathway and OS and RFS by sex

| **Variable^a^** | **OS** | | | | **RFS** | | | |
| --- | --- | --- | --- | --- | --- | --- | --- | --- |
|  | **Females** | | **Males** | | **Females** | | **Males** | |
|  | **HR (95% CI)** | ***P* value** | **HR (95% CI)** | ***P* value** | **HR (95% CI)** | ***P* value** | **HR (95% CI)** | ***P* value** |
| Ribulose 5-phosphate | 1.30 (0.72, 2.35) | 0.388 | 1.09 (0.70, 1.69) | 0.700 | 1.87 (0.96, 3.65) | 0.068 | 1.13 (0.63, 2.03) | 0.673 |
| Lactate | 1.63 (0.45, 5.98) | 0.460 | 1.02 (0.45, 2.34) | 0.960 | 1.17 (0.26, 5.18) | 0.839 | 0.92 (0.28, 3.00) | 0.885 |
| DHAPorG3P | 2.20 (0.75, 6.48) | 0.153 | 0.91 (0.48, 1.75) | 0.785 | 1.35 (0.51, 3.56) | 0.550 | 0.94 (0.45, 1.98) | 0.879 |
| Glucose 6-phosphate | 0.93 (0.48, 1.81) | 0.842 | 0.86 (0.51, 1.43) | 0.553 | 1.90 (1.00, 3.58) | 0.048 | 0.74 (0.44, 1.24) | 0.254 |
| Fructose 6-phosphate | 0.77 (0.32, 1.87) | 0.570 | 0.96 (0.50, 1.82) | 0.890 | 0.26 (0.11, 0.64) | 0.003 | 0.72 (0.37, 1.42) | 0.344 |
| Glycerol 3-phosphate | 2.94 (0.86, 9.99) | 0.084 | 0.89 (0.42, 1.88) | 0.760 | 0.79 (0.27, 2.28) | 0.664 | 0.27 (0.08, 0.88) | 0.029 |
| Phosphoenolpyruvate | 1.16 (0.69, 1.93) | 0.572 | 0.96 (0.70, 1.33) | 0.817 | 1.54 (0.85, 2.77) | 0.154 | 0.98 (0.63, 1.53) | 0.940 |
| Anatomic location = RCC (ref: LCC) | 1.01 (0.27, 3.80) | 0.993 | 0.58 (0.17, 1.91) | 0.367 | 1.03 (0.27, 3.94) | 0.968 | 0.80 (0.23, 2.78) | 0.728 |
| Clinical stage = late (ref: early) | 13.00 (1.96, 86.35) | 0.008 | 3.36 (0.87, 13.02) | 0.080 | 2.00 (0.20, 19.67) | 0.551 | 0.99 (0.18, 5.34) | 0.994 |
| Chemotherapy = Yes (ref: No) | 0.75 (0.13, 4.43) | 0.753 | 0.99 (0.23, 4.20) | 0.987 | 1.65 (0.16, 16.89) | 0.675 | 1.21 (0.24, 6.12) | 0.816 |
| Age | 1.08 (0.98, 1.20) | 0.132 | 1.12 (1.05, 1.19) | 0.001 | 1.01 (0.92, 1.10) | 0.832 | 0.92 (0.84, 1.00) | 0.064 |

^a^ All the variables listed were included in one multivariate Cox PH model. The abundance of each metabolite was treated as a continuous variable and was log_2_ transformed.

**S9 Table.** Multivariate analysis of association between metabolites in lysophospholipids synthesis and OS and RFS for all patients

| **Variable^a^** | **OS** | | | **RFS** | | |
| --- | --- | --- | --- | --- | --- | --- |
|  | **HR (95% CI)** | ***P* value^a^** | **Int. Sex *P* value^b^** | **HR (95% CI)** | ***P* value^a^** | **Int. Sex *P* value^b^** |
| Oleic acid | 1.13 (0.82, 1.56) | 0.457 | 0.271 | 1.33 (0.94, 1.89) | 0.107 | 0.472 |
| L-Palmitoylcarnitine | 1.25 (0.76, 2.06) | 0.378 | 0.784 | 2.64 (1.43, 4.86) | 0.002 | 0.481 |
| Stearoylcarnitine | 0.84 (0.64, 1.09) | 0.190 | 0.900 | 0.76 (0.56, 1.04) | 0.089 | 0.595 |
| Carnitine | 0.52 (0.22, 1.23) | 0.137 | 0.681 | 0.29 (0.11, 0.78) | 0.014 | 0.098 |
| LysoPC(16:0) | 1.22 (0.83, 1.80) | 0.306 | 0.002 | 0.87 (0.44, 1.72) | 0.691 | 0.233 |
| LysoPC(16:1) | 0.68 (0.39, 1.20) | 0.183 | 0.153 | 1.18 (0.61, 2.28) | 0.626 | 0.376 |
| LysoPC(18:1) | 0.75 (0.40, 1.41) | 0.364 | 0.018 | 1.07 (0.53, 2.14) | 0.854 | 0.447 |
| LysoPE(16:0) | 1.66 (0.69, 3.99) | 0.253 | 0.088 | 4.38 (1.75, 10.97) | 0.002 | 0.443 |
| LysoPE(16:1) | 0.98 (0.56, 1.72) | 0.940 | 0.675 | 0.70 (0.33, 1.49) | 0.358 | 0.720 |
| LysoPE(18:0) | 0.84 (0.37, 1.90) | 0.674 | 0.027 | 1.04 (0.47, 2.29) | 0.924 | 0.283 |
| LysoPE(18:1) | 0.88 (0.31, 2.50) | 0.817 | 0.275 | 0.36 (0.13, 1.00) | 0.051 | 0.595 |
| LysoPE(20:1) | 1.08 (0.70, 1.65) | 0.740 | 0.205 | 1.04 (0.64, 1.69) | 0.875 | 0.334 |
| LysoPE(22:5) | 0.88 (0.41, 1.93) | 0.758 | 0.042 | 0.50 (0.24, 1.07) | 0.073 | 0.052 |
| LysoPE(18:2) | 1.52 (0.84, 2.77) | 0.167 | 0.467 | 0.99 (0.49, 1.98) | 0.970 | 0.395 |
| Sex = Male (ref: Female) | 2.30 (1.07, 4.92) | 0.032 | - | 1.58 (0.73, 3.43) | 0.250 | - |
| Anatomic location = RCC (ref: LCC) | 0.59 (0.26, 1.35) | 0.213 | - | 0.74 (0.28, 1.92) | 0.532 | - |
| Clinical stage = late (ref: early) | 4.02 (1.42, 11.38) | 0.009 | - | 3.70 (1.26, 10.84) | 0.017 | - |
| Chemotherapy = Yes (ref: No) | 1.42 (0.48, 4.16) | 0.523 | - | 0.91 (0.30, 2.72) | 0.862 | - |
| Age | 1.11 (1.06, 1.16) | <0.001 | - | 0.96 (0.90, 1.02) | 0.174 | - |

Abbreviation: LysoPC, lysophosphatidylcholine; LysoPE, lysophosphatidylethanolamine.

^a^ All the variables listed were included in one multivariate Cox PH model where the abundance of each metabolite was treated as a continuous variable and was log_2_ transformed.

^b^ *P* value of the interaction between the metabolite and sex.

**S10 Table.** Multivariate analysis of association between metabolites in lysophospholipids synthesis and OS and RFS by sex

| **Variable^a^** | **OS** | | | | **RFS** | | | |
| --- | --- | --- | --- | --- | --- | --- | --- | --- |
|  | **Females** | | **Males** | | **Females** | | **Males** | |
|  | **HR (95% CI)** | ***P* value** | **HR (95% CI)** | ***P* value** | **HR (95% CI)** | ***P* value** | **HR (95% CI)** | ***P* value** |
| Oleic acid | 1.05 (0.50, 2.20) | 0.905 | 1.66 (1.02, 2.69) | 0.041 | 2.27 (0.95, 5.44) | 0.065 | 0.91 (0.53, 1.57) | 0.728 |
| L-Palmitoylcarnitine | 1.55 (0.61, 3.89) | 0.355 | 2.99 (1.03, 8.69) | 0.044 | 3.93 (1.29, 12.04) | 0.016 | 1.79 (0.57, 5.64) | 0.323 |
| Stearoylcarnitine | 0.65 (0.41, 1.05) | 0.076 | 0.60 (0.34, 1.05) | 0.074 | 0.60 (0.37, 0.99) | 0.047 | 0.75 (0.42, 1.34) | 0.329 |
| Carnitine | 6.83 (0.11, 431.17) | 0.363 | 0.19 (0.06, 0.67) | 0.009 | 5.84 (0.08, 435.95) | 0.422 | 0.19 (0.03, 1.03) | 0.054 |
| LysoPC(16:0) | 2.07 (1.06, 4.07) | 0.034 | 0.64 (0.27, 1.54) | 0.319 | 1.06 (0.37, 3.04) | 0.920 | 0.43 (0.08, 2.21) | 0.313 |
| LysoPC(16:1) | 0.39 (0.11, 1.37) | 0.144 | 0.52 (0.22, 1.27) | 0.151 | 2.71 (0.66, 11.17) | 0.169 | 0.86 (0.27, 2.70) | 0.798 |
| LysoPC(18:1) | 1.56 (0.43, 5.75) | 0.501 | 0.29 (0.08, 1.00) | 0.050 | 0.41 (0.10, 1.65) | 0.210 | 7.82 (1.30, 47.16) | 0.025 |
| LysoPE(16:0) | 1.66 (0.35, 7.95) | 0.524 | 12.72 (2.20, 73.55) | 0.005 | 1.52 (0.34, 6.83) | 0.588 | 6.58 (0.99, 43.95) | 0.052 |
| LysoPE(16:1) | 1.27 (0.38, 4.30) | 0.695 | 1.16 (0.50, 2.73) | 0.726 | 1.07 (0.26, 4.45) | 0.927 | 0.70 (0.18, 2.73) | 0.604 |
| LysoPE(18:0) | 2.56 (0.65, 10.11) | 0.179 | 0.10 (0.02, 0.44) | 0.002 | 4.21 (0.79, 22.39) | 0.092 | 2.64 (0.36, 19.31) | 0.338 |
| LysoPE(18:1) | 0.57 (0.06, 5.17) | 0.618 | 0.80 (0.14, 4.63) | 0.806 | 0.07 (0.01, 0.63) | 0.017 | 0.30 (0.04, 2.19) | 0.233 |
| LysoPE(20:1) | 0.71 (0.25, 2.04) | 0.521 | 1.14 (0.62, 2.11) | 0.679 | 0.33 (0.07, 1.47) | 0.147 | 1.37 (0.61, 3.06) | 0.444 |
| LysoPE(22:5) | 0.78 (0.21, 2.93) | 0.714 | 2.47 (0.66, 9.16) | 0.179 | 5.38 (1.02, 28.42) | 0.048 | 0.10 (0.02, 0.62) | 0.013 |
| LysoPE(18:2) | 1.06 (0.20, 5.45) | 0.948 | 2.41 (1.12, 5.20) | 0.024 | 1.59 (0.31, 8.26) | 0.580 | 0.50 (0.14, 1.77) | 0.286 |
| Anatomic location = RCC (ref: LCC) | 1.05 (0.29, 3.79) | 0.941 | 0.09 (0.02, 0.43) | 0.002 | 1.11 (0.19, 6.58) | 0.909 | 1.00 (0.14, 6.97) | 1.000 |
| Clinical stage = late (ref: early) | 18.99 (2.31, 155.90) | 0.006 | 15.45 (2.48, 96.39) | 0.003 | 4.40 (0.24, 79.55) | 0.316 | 2.49 (0.36, 17.33) | 0.355 |
| Chemotherapy = Yes (ref: No) | 0.84 (0.13, 5.37) | 0.853 | 0.83 (0.16, 4.37) | 0.827 | 2.24 (0.14, 36.07) | 0.570 | 0.51 (0.08, 3.33) | 0.485 |
| Age | 1.10 (1.01, 1.21) | 0.038 | 1.30 (1.16, 1.45) | <0.001 | 1.05 (0.96, 1.16) | 0.278 | 0.86 (0.76, 0.97) | 0.018 |

Abbreviation: LysoPC, lysophosphatidylcholine; LysoPE, lysophosphatidylethanolamine.

^a^ All the variables listed were included in one multivariate Cox PH model. The abundance of each metabolite was treated as a continuous variable and was log_2_ transformed.

**S11 Table.** Multivariate analysis of association between metabolites in methionine metabolism and OS and RFS for all patients

| **Variable^a^** | **OS** | | | **RFS** | | |
| --- | --- | --- | --- | --- | --- | --- |
|  | **HR (95% CI)** | ***P* value^a^** | **Int. Sex *P* value^b^** | **HR (95% CI)** | ***P* value^a^** | **Int. Sex *P* value^b^** |
| Dimethylglycine | 0.99 (0.64, 1.52) | 0.955 | 0.390 | 0.74 (0.46, 1.20) | 0.228 | 0.555 |
| Methionine | 1.26 (0.73, 2.19) | 0.408 | 0.045 | 1.90 (1.01, 3.57) | 0.045 | 0.022 |
| S-adenosylhomocysteine (SAH) | 0.96 (0.75, 1.23) | 0.751 | 0.078 | 0.96 (0.72, 1.27) | 0.753 | 0.931 |
| S-adenosylmethionine (SAM) | 1.08 (0.88, 1.31) | 0.467 | 0.355 | 1.07 (0.86, 1.32) | 0.541 | 0.765 |
| Serine | 0.56 (0.34, 0.92) | 0.023 | 0.039 | 0.49 (0.29, 0.83) | 0.008 | 0.045 |
| Sex = Male (ref: Female) | 2.02 (0.99, 4.13) | 0.054 | - | 1.14 (0.53, 2.44) | 0.740 | - |
| Anatomic location = RCC (ref: LCC) | 0.77 (0.38, 1.58) | 0.483 | - | 0.62 (0.28, 1.37) | 0.235 | - |
| Clinical stage = late (ref: early) | 5.32 (2.01, 14.11) | 0.001 | - | 1.88 (0.60, 5.85) | 0.277 | - |
| Chemotherapy = Yes (ref: No) | 1.02 (0.40, 2.61) | 0.962 | - | 1.24 (0.41, 3.77) | 0.703 | - |
| Age | 1.10 (1.05, 1.15) | <0.001 | - | 0.96 (0.91, 1.02) | 0.189 | - |

^a^ All the variables listed were included in one multivariate Cox PH model where the abundance of each metabolite was treated as a continuous variable and was log_2_ transformed.

^b^ *P* value of the interaction between the metabolite and sex.

**S12 Table.** Multivariate analysis of association between metabolites in methionine metabolism and OS and RFS by sex

| **Variable^a^** | **OS** | | | | **RFS** | | | |
| --- | --- | --- | --- | --- | --- | --- | --- | --- |
|  | **Females** | | **Males** | | **Females** | | **Males** | |
|  | **HR (95% CI)** | ***P* value** | **HR (95% CI)** | ***P* value** | **HR (95% CI)** | ***P* value** | **HR (95% CI)** | ***P* value** |
| Dimethylglycine | 0.69 (0.33, 1.42) | 0.308 | 1.45 (0.77, 2.74) | 0.250 | 0.63 (0.29, 1.36) | 0.239 | 0.69 (0.33, 1.46) | 0.331 |
| Methionine | 2.52 (0.27, 23.62) | 0.419 | 1.30 (0.64, 2.65) | 0.465 | 5.49 (0.49, 61.80) | 0.168 | 1.39 (0.62, 3.10) | 0.423 |
| S-adenosylhomocysteine (SAH) | 0.87 (0.56, 1.36) | 0.545 | 1.09 (0.76, 1.58) | 0.631 | 1.07 (0.67, 1.72) | 0.776 | 0.81 (0.54, 1.21) | 0.304 |
| S-adenosylmethionine (SAM) | 1.18 (0.78, 1.78) | 0.426 | 1.04 (0.80, 1.36) | 0.757 | 1.08 (0.72, 1.62) | 0.709 | 1.20 (0.90, 1.60) | 0.211 |
| Serine | 0.48 (0.05, 4.59) | 0.527 | 0.37 (0.19, 0.72) | 0.004 | 0.31 (0.03, 3.13) | 0.322 | 0.49 (0.24, 0.98) | 0.044 |
| Anatomic location = RCC (ref: LCC) | 1.32 (0.42, 4.17) | 0.636 | 0.58 (0.22, 1.58) | 0.289 | 0.81 (0.26, 2.53) | 0.714 | 0.43 (0.12, 1.50) | 0.184 |
| Clinical stage = late (ref: early) | 9.44 (1.44, 61.68) | 0.019 | 9.11 (2.22, 37.39) | 0.002 | 2.25 (0.42, 12.14) | 0.345 | 0.76 (0.13, 4.39) | 0.764 |
| Chemotherapy = Yes (ref: No) | 1.12 (0.24, 5.18) | 0.886 | 0.81 (0.23, 2.82) | 0.742 | 2.04 (0.39, 10.71) | 0.398 | 1.67 (0.34, 8.30) | 0.529 |
| Age | 1.06 (0.98, 1.14) | 0.134 | 1.16 (1.08, 1.25) | <0.001 | 0.97 (0.90, 1.06) | 0.541 | 0.93 (0.86, 1.01) | 0.076 |

^a^ All the variables listed were included in one multivariate Cox PH model. The abundance of each metabolite was treated as a continuous variable and was log_2_ transformed.

**S13 Table.** Multivariate analysis of association between metabolites in polyamine synthesis and OS and RFS for all patients

| **Variable^a^** | **OS** | | | **RFS** | | |
| --- | --- | --- | --- | --- | --- | --- |
|  | **HR (95% CI)** | ***P* value^a^** | **Int. Sex *P* value^b^** | **HR (95% CI)** | ***P* value^a^** | **Int. Sex *P* value^b^** |
| *N*^1^, *N*^12^-diacetylspermine (DAS) | 0.87 (0.62, 1.23) | 0.439 | 0.356 | 1.01 (0.68, 1.50) | 0.954 | 0.336 |
| Spermidine | 1.30 (0.64, 2.63) | 0.465 | 0.594 | 0.72 (0.34, 1.53) | 0.396 | 0.329 |
| Spermine | 1.16 (0.83, 1.62) | 0.386 | 0.082 | 1.06 (0.76, 1.49) | 0.719 | 0.933 |
| N^1^-acetylspermine | 1.07 (0.78, 1.48) | 0.675 | 0.887 | 1.24 (0.89, 1.74) | 0.203 | 0.109 |
| Ornithine | 0.97 (0.54, 1.74) | 0.927 | 0.105 | 0.84 (0.51, 1.39) | 0.503 | 0.206 |
| Arginine | 1.38 (0.83, 2.27) | 0.210 | 0.070 | 2.03 (1.14, 3.60) | 0.016 | 0.037 |
| Citrulline | 0.96 (0.69, 1.34) | 0.818 | 0.002 | 1.05 (0.73, 1.51) | 0.801 | 0.023 |
| Argininosuccinic acid | 0.69 (0.48, 0.98) | 0.039 | 0.123 | 0.72 (0.49, 1.04) | 0.081 | 0.055 |
| Sex = Male (ref: Female) | 2.15 (1.08, 4.29) | 0.030 | - | 1.17 (0.54, 2.56) | 0.690 | - |
| Anatomic location = RCC (ref: LCC) | 0.81 (0.39, 1.71) | 0.584 | - | 0.65 (0.29, 1.48) | 0.306 | - |
| Clinical stage = late (ref: early) | 6.11 (1.91, 19.57) | 0.002 | - | 1.93 (0.64, 5.81) | 0.242 | - |
| Chemotherapy = Yes (ref: No) | 0.92 (0.28, 2.95) | 0.882 | - | 1.14 (0.36, 3.63) | 0.824 | - |
| Age | 1.10 (1.05, 1.15) | <0.001 | - | 0.96 (0.90, 1.01) | 0.120 | - |

^a^ All the variables listed were included in one multivariate Cox PH model where the abundance of each metabolite was treated as a continuous variable and was log_2_ transformed.

^b^ *P* value of the interaction between the metabolite and sex.

**S14 Table.** Multivariate analysis of association between metabolites in polyamine synthesis and OS and RFS by sex

| **Variable^a^** | **OS** | | | | **RFS** | | | |
| --- | --- | --- | --- | --- | --- | --- | --- | --- |
|  | **Females** | | **Males** | | **Females** | | **Males** | |
|  | **HR (95% CI)** | ***P* value** | **HR (95% CI)** | ***P* value** | **HR (95% CI)** | ***P* value** | **HR (95% CI)** | ***P* value** |
| N^1^, N^12^-diacetylspermine (DAS) | 2.45 (0.96, 6.22) | 0.059 | 0.66 (0.42, 1.02) | 0.060 | 1.10 (0.50, 2.42) | 0.813 | 1.07 (0.61, 1.90) | 0.812 |
| Spermidine | 0.51 (0.15, 1.67) | 0.264 | 3.21 (1.04, 9.93) | 0.043 | 0.28 (0.09, 0.91) | 0.034 | 4.18 (1.03, 16.96) | 0.045 |
| Spermine | 3.88 (1.50, 10.06) | 0.005 | 0.71 (0.43, 1.18) | 0.184 | 1.61 (0.90, 2.88) | 0.106 | 0.56 (0.32, 1.00) | 0.048 |
| N^1^-acetylspermine | 0.37 (0.16, 0.86) | 0.020 | 1.14 (0.74, 1.75) | 0.557 | 0.92 (0.46, 1.83) | 0.816 | 1.62 (0.99, 2.63) | 0.054 |
| Ornithine | 0.64 (0.10, 4.28) | 0.646 | 0.78 (0.40, 1.55) | 0.481 | 0.41 (0.10, 1.67) | 0.213 | 0.86 (0.45, 1.67) | 0.660 |
| Arginine | 5.63 (1.33, 23.78) | 0.019 | 1.06 (0.54, 2.08) | 0.869 | 3.22 (1.14, 9.13) | 0.028 | 1.57 (0.74, 3.34) | 0.237 |
| Citrulline | 2.57 (1.04, 6.33) | 0.040 | 0.73 (0.47, 1.15) | 0.178 | 1.24 (0.60, 2.59) | 0.561 | 0.88 (0.51, 1.50) | 0.630 |
| Argininosuccinic acid | 0.57 (0.23, 1.38) | 0.212 | 0.74 (0.47, 1.19) | 0.214 | 1.20 (0.53, 2.73) | 0.667 | 0.43 (0.24, 0.78) | 0.005 |
| Anatomic location = RCC (ref: LCC) | 0.33 (0.06, 1.69) | 0.182 | 0.80 (0.31, 2.08) | 0.648 | 0.55 (0.14, 2.08) | 0.375 | 0.50 (0.15, 1.69) | 0.265 |
| Clinical stage = late (ref: early) | 119.96 (4.62, 3116.09) | 0.004 | 3.56 (0.94, 13.45) | 0.061 | 1.27 (0.19, 8.70) | 0.805 | 0.95 (0.20, 4.60) | 0.947 |
| Chemotherapy = Yes (ref: No) | 0.10 (0.01, 1.50) | 0.096 | 2.20 (0.56, 8.66) | 0.258 | 4.54 (0.59, 34.72) | 0.145 | 1.14 (0.21, 6.14) | 0.878 |
| Age | 1.18 (1.05, 1.33) | 0.007 | 1.15 (1.07, 1.23) | <0.001 | 0.97 (0.87, 1.08) | 0.625 | 0.90 (0.82, 0.98) | 0.014 |

^a^ All the variables listed were included in one multivariate Cox PH model. The abundance of each metabolite was treated as a continuous variable and was log_2_ transformed.
